# From SARS and MERS to COVID-19: a review of the quality and responsiveness of clinical management guidelines in outbreak settings

**DOI:** 10.1101/2021.01.12.21249654

**Authors:** Samuel Lipworth, Ishmeala Rigby, Vincent Cheng, Peter Bannister, Eli Harriss, Karen Cook, Erhui Cai, Mais Tattan, Terrence Epie, Lakshmi Manoharan, Kate Lambe, Melina Michelen, Anna Vila Gilibets, Andrew Dagens, Louise Sigfrid, Peter Horby

## Abstract

**Objective:** To assess the responsiveness and quality of clinical management guidelines (CMGs) in SARS, MERS and COVID-19 and determine whether this has improved over time.

**Design:** Rapid literature review, quality assessment and focus group consultation.

**Data Sources:** – Google and Google Scholar were systematically searched from inception to 6^th^ June 2020.This was supplemented with hand searches of national and international public health agency and infectious disease society websites as well as directly approaching clinical networks in regions where few CMGs had been identified via the primary search.

**Eligibility Criteria:** CMGs for the treatment of COVID-19/SARS/MERS providing recommendations on supportive care and/or specific treatment.

**Methods:** Data extraction was performed using a standardised form. The Appraisal of Guidelines for Research and Evaluation (AGREE-II) tool was used to evaluate the quality of the CMGs. Six COVID-19 treatments were selected to assess the responsiveness of a subset of guidelines and their updates to 20^th^ November 2020. We ran two sessions of focus groups with patient advocates to elicit their views on guideline development.

**Results:** We included 37 COVID-19, six SARS, and four MERS CMGs. Evidence appraisals in CMGs generally focused on novel drugs rather than basic supportive care; where evidence for the latter was provided it was generally of a low quality. Most CMGs had major methodological flaws (only two MERS-CoV and four COVID-19 CMGs were recommended for use by both reviewers without modification) and there was no evidence of improvement in quality over time. CMGs scored lowest in the following AGREE-II domains: scope and purpose, editorial independence, stakeholder engagement, and rigour of development. Of the COVID-19 CMGs, only eight included specific guidance for the management of elderly patients and only ten for high-risk groups; a further eight did not specify the target patient group at all. Early in the pandemic, multiple guidelines recommended unproven treatments and whilst in general findings of major clinical trials were eventually adopted, this was not universally the case. Eight guidelines recommended that use of unproven agents should be considered on a case-by-case basis. Patient representatives expressed concern about the lack of engagement with them in CMG development and that these documents are not accessible to non-experts.

**Conclusion:** The quality of most CMGs produced in coronaviridae outbreaks is poor and we have found no evidence of improvement over time, highlighting that current development frameworks must be improved. There is an need to strengthen the evidence base surrounding basic supportive care and develop methods to engage patients in CMG development from the beginning in outbreak settings.

**Systematic review registration:** PROSPERO CRD42020167361

## Introduction

Clinical management guidelines (CMGs) are useful tools to help clinicians provide quality, evidence-based care to patients. Their utility is potentially even greater in an outbreak setting when clinicians are faced with the challenges of managing a new pathogen combined with increased pressures on healthcare services and redeployment to areas in which they have limited experience. Outbreaks are however also associated with significant time pressure and high levels of uncertainty, making production of methodologically rigorous guidelines difficult.^1^

The SARS-CoV-2 pandemic has highlighted the disproportionate impact of infectious disease outbreaks on vulnerable (e.g. the elderly and immunosuppressed)^2^ and socioeconomically disadvantaged groups in society.^3^ Infectious diseases often present differently in these populations and yet most CMGs produced early in the pandemic did not provide specific advice for the management of these groups.^1^ As knowledge about new diseases increases as time elapses, the inclusivity, quality and usefulness of CMGs should also improve. Pandemics such as COVID-19 are likely to occur with increasing frequency throughout the 21st Century and a failure to improve the processes by which clinical practice learns and responds will ultimately lead to unnecessary morbidity and mortality.^4^

In this manuscript, we track the evolution of clinical management guidelines across three coronaviridae pan/epidemics: SARS-CoV-1, MERS-CoV and SARS-CoV-2. We particularly focus on whether the rigour of development of guidelines and inclusivity of vulnerable groups has improved between these outbreaks and over the course of the COVID-19 pandemic. We aim to identify the strengths and weaknesses of guidelines produced in these settings and to evaluate whether lessons from previous outbreaks have been learnt. For a subset of guidelines in the current SARS-CoV-2 pandemic we also examine how responsive these CMGs are in incorporating new evidence from the latest clinical trials. In doing so we ask the bigger question of how clinical management guidelines can be improved as health professionals continue to manage large numbers of COVID-19 patients and for future pandemics.

## Methods

This review is an update of a rapid review^1^ and part of a wider project evaluating the availability, quality and inclusivity of clinical management guidelines for high consequence infectious diseases (HCID). The Preferred Reporting in Systematic Reviews and Meta-Analyses (PRISMA) checklist was used to construct this review (Figure 1).^5^

**Figure 1:**
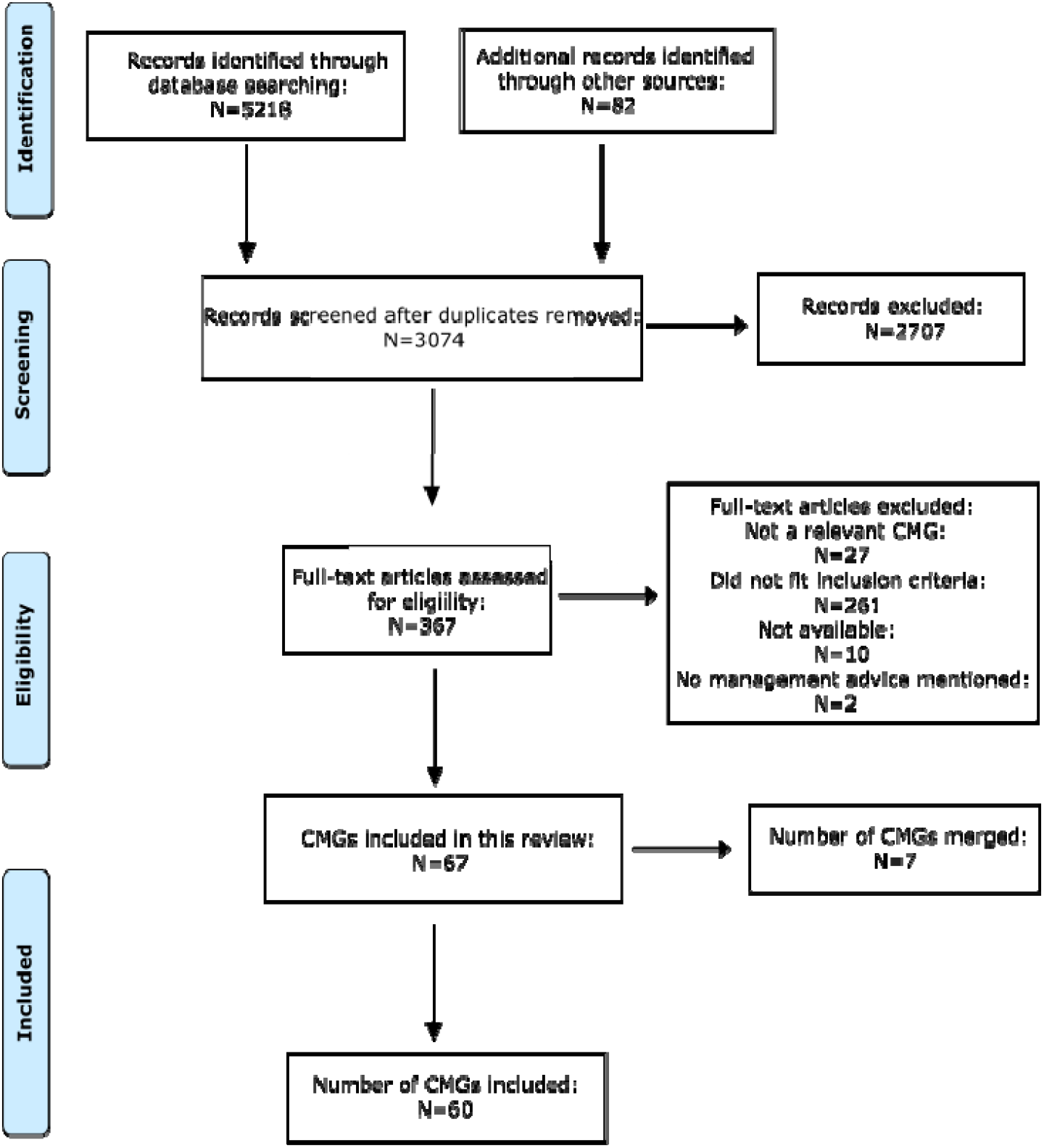
PRISMA diagram.

### Search strategy

In a previous review, we found that most CMGs were not published in peer-reviewed journals and rarely indexed in the electronic databases.^1^ We therefore focussed our efforts on extensive hand-searches of the grey literature using a combination of systematic Google and Google Scholar searches and by specifically searching Ministry of Health, national public health agency institutions, World Health Organisation (WHO), Centres for Disease Control and other infectious disease society websites with pre-defined keywords (supplement). This was complemented by utilising the International Severe Acute Respiratory and Emerging Infection Consortium (ISARIC)^6^ network to contact clinical networks in regions where limited numbers of CMGs where initially identified. Finally, we searched reference lists of included CMGs. We aimed to identify a globally representative sample of CMGs, focusing on international and national CMGs in this review for feasibility and because these likely inform the development of locally developed guidelines at a hospital/regional level. The full search strategy is shown in the Supplementary Methods. The search was completed on the 6^th^ June 2020.

### Inclusion/exclusion criteria

COVID-19, SARS, and MERS CMGs that included recommendations intended to optimize patient care were included.^7^ Guidelines that were substantially local policy documents or focused primarily on infection control/diagnostics were therefore excluded. There were no language restrictions.

### Screening

Records identified from searches were independently screened, first by title and abstract, followed by full text, by two reviewers. Individuals with knowledge of the language the CMG was written in were used; where this was not possible translations were produced with Google Translate. Any disagreements were resolved by a third reviewer.

### Data extraction

We utilised a standardised form to extract data (supplement). Data was extracted by one reviewer and checked by a second reviewer.

### Quality assessment

The quality of each CMG was assessed using the Appraisal of Guidelines, Research and Evaluation version II (AGREE-II).^8^ The tool consists of 23 questions (scored on a 7-point scale, from 1 (strongly disagree) to 7 (strongly agree)) across six key domains (scope and purpose; stakeholder involvement; rigour of development; clarity of presentation; applicability; editorial independence). All CMGs were assessed using AGREE-II by two reviewers independently.

CMGs where there was significant discordance in the reviewers assessments were identified by calculating Cohen’s Kappa; a threshold of 0.4 was used to trigger further discussion between reviewers to resolve major disagreements. We considered three measures of whether a CMG was high quality: an overall weighted score ≥ 0.7 (threshold suggested by the AGREE-II developers), weighted score ≥ 0.7 on domains 3 and 5 (rigour of development and applicability, previously shown to be most predictive of overall score^9^) and reviewers’ overall assessment of whether they would recommend use of the CMG. Weighted scores were calculated according to the formula presented the AGREE-II manual8 : 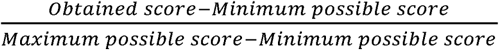

### Responsiveness/Quality over time (subset analysis)

We tracked a subset of 11 COVID-19 CMGs (selected because they also featured in our earlier rapid review ^1^) over time to assess their responsiveness to key results from randomised clinical trials (RCTs) for six treatments (hydroxychloroquine, convalescent plasma, lopinavir-ritonavir, remdesivir, dexamethasone and tocilizumab). For each CMG in this subset, we also compared the AGREE-II scores to those given to earlier versions at the beginning of the pandemic in our previous review.

### Patient and Public Involvement

Members of the public were invited to comment on the results and interpretation of our study via a social media COVID research participation group. Two semi-structured focus groups, facilitated by two authors, were held via a teleconference call. Participants worked with review authors to comment on the methodology and inform the interpretation and presentation of results.

### Statistics

Statistical analysis was performed in the R language for statistical computing^10^ version 4.0.2 with the ggplot library used to produce graphics.^11^

## Results

In the main searches completed on 6^th^ June 2020, we identified 47 CMGs (Figure 1). 37 covered clinical management of COVID-19, four of MERS and six of SARS. Most COVID-19 CMGs were developed by government agencies and published on the websites as standalone documents or acquired via the ISARIC^6^ network. In contrast, SARS and MERS guidelines were generally published in peer-reviewed journals. Although we attempted to ensure that there were at least five national COVID-19 CMGs per continent, we found fewer guidelines produced in Australasia (n=1), South America (n=3) and Africa (n=6), compared to North America (n=7), Europe (n=12), and Asia (n=15). By the World Bank definition,^12^ most guidelines were produced in high (n=21) and upper middle (n=14), followed by lower middle (n=8) and low-income countries (n=1). Three CMGs where produced by international agencies.^13-15^

### Quality evaluation

Most CMGs were not high quality by any of the three objective measures we used. For example, (27%) 10/37 of COVID-19 CMGs had an overall score of 0.7 or above compared to 2/4 (50%) MERS and 0/6 (0%) SARS. Only one guideline scored 0.7 or more for domains 3 (clarity of presentation) and 5 (rigour of development) (Korean Society of Infectious Disease MERS-CoV guideline^16^); notably no COVID-19 guidelines met this standard. In total 25/47 CMGs were recommended for use by both reviewers though there were only six (two MERS-CoV and four COVID-19) where both reviewers agreed no modification was desirable. The highest score of these were COVID-19 CMGs developed by the Infectious Diseases Society of America (IDSA),^17^ Surviving Sepsis Campaign,^14^ and a MERS CMG developed by the Korean Society of Infectious Disease.^16^ These were notable for their clear expression of clinical questions which were answered rigorously using a defined methodology and were presented to a high standard.

Considering all included CMGs, quality was not equal across the domains of the AGREE-II tool (Kruskall-Wallis p<0.001) and there was a wide distribution of scores within domains (Figure 2). Editorial independence’ (median weighted score 0 interquartile range (IQR) 0-0.08) and rigour of development (median weighted score 0.23 (IQR 0.13-0.35) were the lowest scoring domains. The low scores for editorial independence were generally because there was no statement about the role of the funding body and many lacked conflicts of interest declarations. Low scores for rigour of development reflected the absence of a description of a systematic evidence search methodology, a lack of explicit links to supporting evidence and unclear methods for selecting key recommendations. CMGs scored better for the ‘Clarity of Presentation’ domain (median weighted score 0.67, IQR (0.47-0.81)). There was weak evidence of a difference in the overall scores of guidelines produced by academic societies vs public health agencies (median 4.5 (IQR 3.5-5.5) vs. median 3.8 (IQR 3.0-4.5), Wilcox p=0.06).

**Figure 2:**
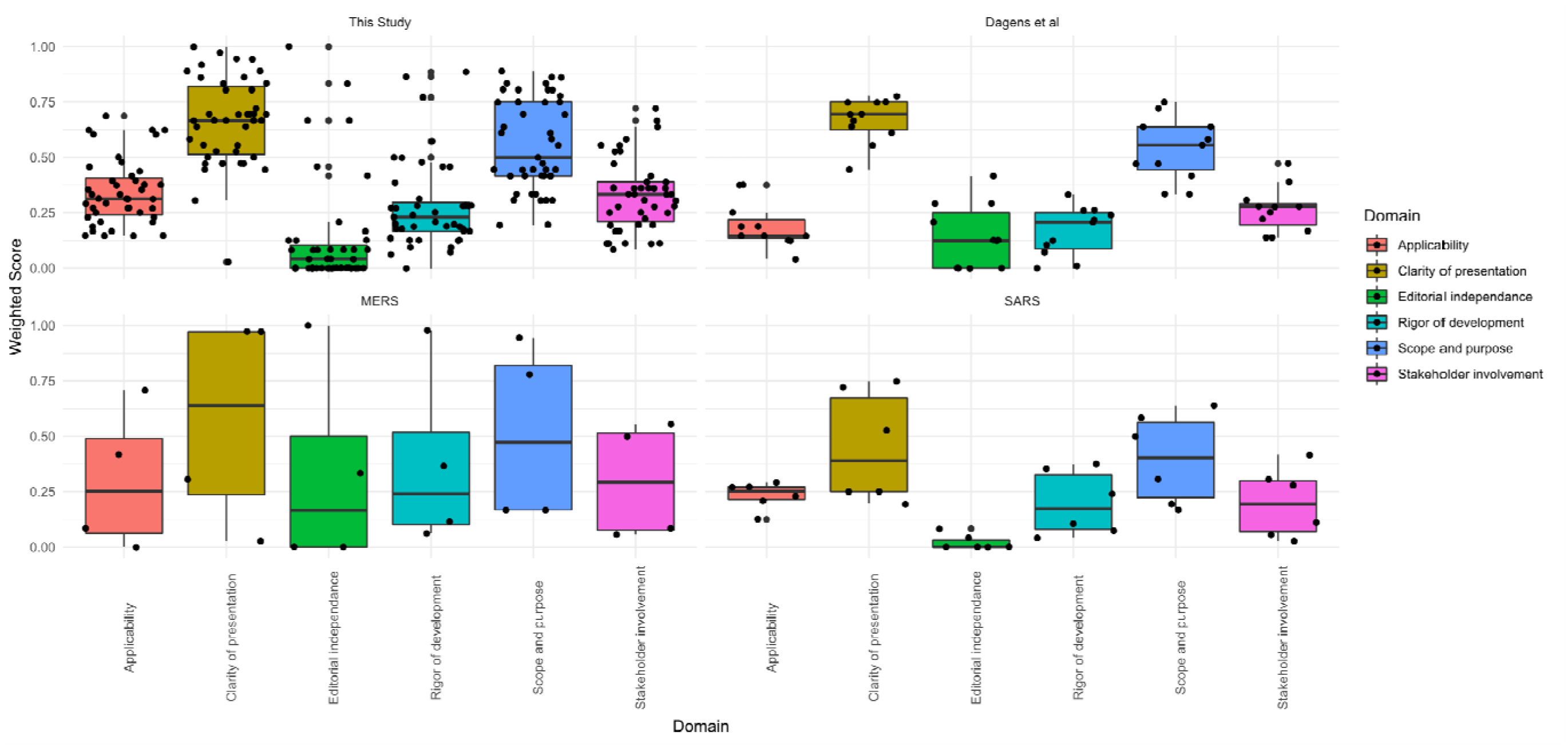
Weighted scores for the six domains of the AGREE-II tool for the four groups of CMGs included. Dagens et al^1^ refers to CMGs published in the early part of the COVID-19 pandemic. The boxplots show median and interquartile range (IQR) with the upper/lower whiskers showing the position of 1.5* IQR; individual datapoints are represented by black dots.

### No improvement in quality over time

To evaluate whether the quality of CMGs improved over time, we appraised CMGs from three coronaviridae outbreaks (SARS 2002-2004, MERS 2012 and COVID-19 2020). There was no evidence that overall scores were different between these outbreaks (SARS median 0.47 (IQR 0.33-0.61), MERS median 0.54 (IQR 0.27-0.83), COVID-19 median 0.57 (IQR 0.50-0.71), Kruskal-Wallis p=0.35). Notably there was also no evidence of improvement in any of the six domains measured by the AGREE-II tool between the initial and updated COVID-19 guidelines (Bonferroni adjusted paired Wilcoxon Rank Sum p>0.1 in all cases, Figure 2).

### Inclusivity of CMGs

Many CMGs were not specific in their description of the target population. This was reflected in the fact that only 34% (16/47) of all CMGs scored five or more in this AGREE-II question. Most guidelines included general advice for the management of adults, pregnant women, and children, but older and other high-risk groups patients (e.g. immunosuppressed) were notable omissions from many guidelines (Table 1). There were however some examples where this was done well for example in the WHO CMG which includes specific sections relating to the care of older people and pregnant women with COVID-19 as well as guidance on palliative care.

### Supportive Care Recommendations

Nearly all CMGs gave recommendations for aspects of basic supportive care though there was generally little or no supporting literature cited. Most suggested target saturations and methods of oxygen delivery in hypoxic patients, but there were often no links to or discussion of relevant studies. For example, the WHO CMG notes that there is no evidence based guidance for the use of high flow nasal cannula (HFNC) in this setting and recommends that selected patients with COVID-19 and mild acute respiratory distress syndrome (ARDS) be considered for a therapeutic trial of Non-invasive ventilation (NIV).^13^ No literature is provided to support this recommendation and the criteria for selecting patients for such a trial are unclear. Similarly the Surviving Sepsis COVID-19 CMG recommends the use of HFNC over NIV but notes that the quality of evidence is weak.^14^ As a further example, (62%) 23/37 COVID-19 guidelines recommended the use of antimicrobial therapy if bacterial superinfection was clinically suspected. However most did not give guidance as to how this decision should be made nor give clear criteria for stopping (table 2). Three guidelines recommended the use of procalcitonin to guide antimicrobial use though did not provide specific thresholds.^18-20^ Some stratified recommendations for initiating antibiotics by severity of presentation.^21-23^

### Recommendations prior to the availability of high-quality evidence

CMGs varied markedly in their approach to uncertainty of therapeutic efficacy. Some noted the presence of ongoing clinical trials but made no comment on whether an agent should be used whilst others explicitly stated that no recommendation either way could be made. There were several instances where CMGs recommended that where such uncertainty existed, individual case-by-case decisions should be made based on clinical judgement (e.g. COREB for remdesivir/hydroxychloroquine/lopinavir-ritonavir^24^ and the Korean Society of Infectious Diseases for Intravenous (IV) immunoglobulin^25^). Others (e.g. US CDC,^26^ IDSA^17^ and WHO^13^) stated that where there was a lack of evidence, agents should only be used in the context of a clinical trial (Figure 3).

**Figure 3:**
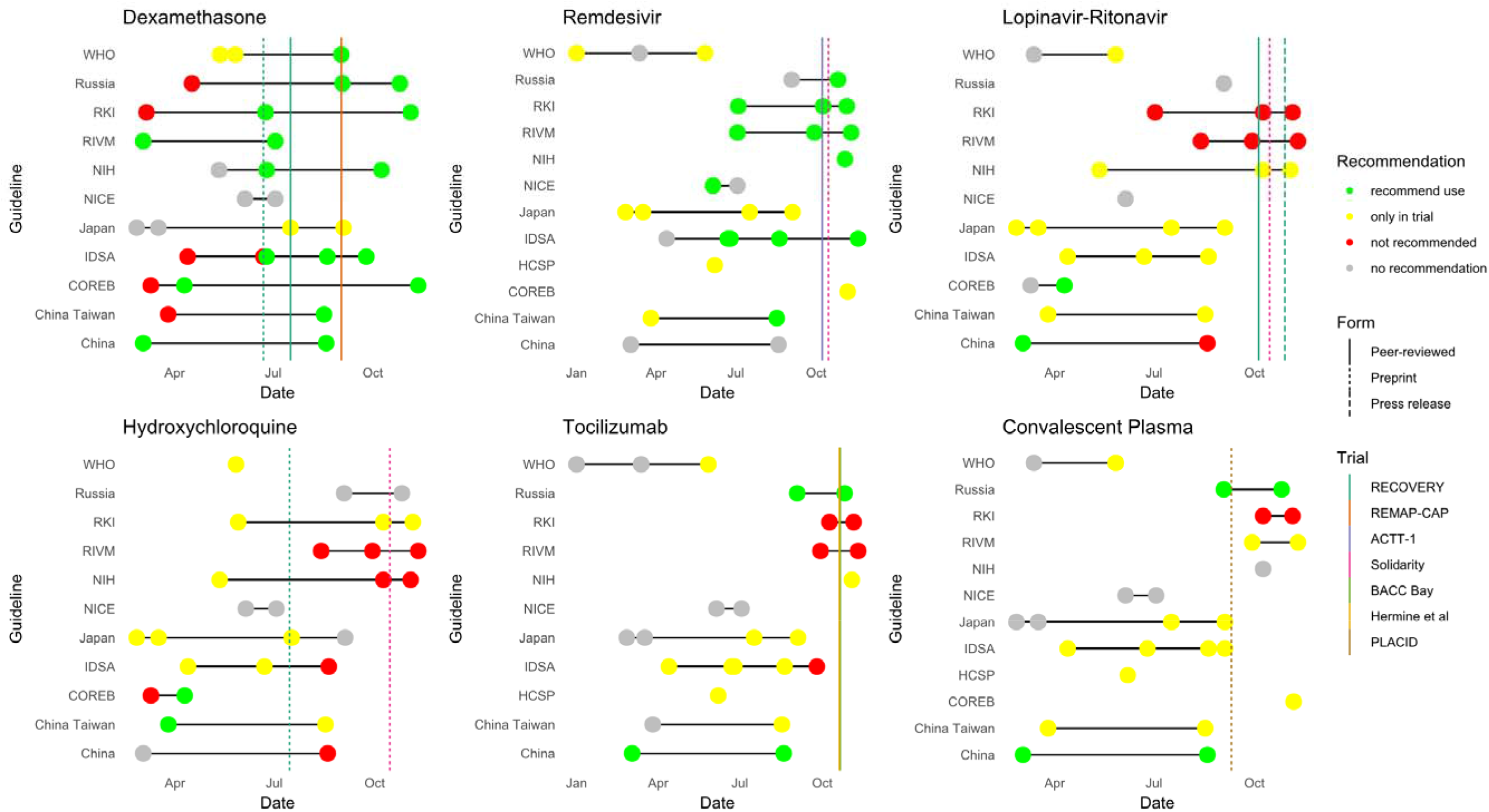
Adoption of evidence from clinical trials by CMGs over time. Intersecting vertical lines show the publication of key clinical trials either as peer- reviewed articles/pre-prints or press-releases. Dots show the publication of CMGs by bodies shown on the y axis coloured according to the recommendation made.

### Responsiveness to emerging evidence

We followed a group of COVID-19 guidelines and tracked their recommendations on six treatments between January and November 2020 (Figure 3). Of the COVID-19 CMGs, 6/11 (55%) changed their guidance on the use of Dexamethasone in response to the results of the Randomised Evaluation of COVID-19 Therapy (RECOVERY) trial and the Embedded, Multi-factorial, Adaptive Platform Trial for Community-Acquired Pneumonia (REMAP-CAP) trial.^17 20 23 26-37^ Four CMGs initially recommended the use of lopinavir/ritonavir and/or hydroxychloroquine, and all except one, a Russian CMG which noted anecdotal success with its use and recommended use in moderate cases^20^, recommended against its use after the publication of the RECOVERY/SOLIDARITY trials,^36 38^ In the case of Remdesivir, 5/11 (45%) CMGs recommended its use prior to the publication of the results of the Adaptive COVID-19 Treatment Trial (ACTT-1).^39^ A similar theme was apparent in the SARS/MERS guidelines where 4/10 (40%) recommended the use of corticosteroids either absolutely or on a case-by-case basis, despite a lack of evidence.^40^

### Stakeholder engagement

In our AGREE-II evaluations, CMGs were consistently poorly rated for their involvement of patient groups in their development (median score 0, (IQR 0-0)). Whilst our patient group acknowledged the need for speed, they unanimously and strongly believed that public involvement in the production of CMGs for COVID-19 would have been desirable to ensure that the patient perspective is incorporated. For example, whilst specialists are understandably focused on acute and critical care, the group felt that patient involvement might have highlited the need for better integration with primary care and the potential utility of ambulatory monitoring (e.g. pulse oximetry). Involving patients early in a pandemic is understandably challenging, nevertheless, development of a pre-identified group that can quickly be available when needed in future was suggested. The patient group were of the opinion that stakeholder involvement, rigour of development and editorial independence (the worst performing domains in our AGREE-II evaluation) were important and that a compromise in their quality was not acceptable despite the mitigating consideration of a pandemic setting. All participants agreed that making CMGs more accessible to a lay audience is something they would value. A few individuals proposed the use of guideline summaries written in plain English, or in the form of infographics and videos. Participants felt that this would better enable patient centred care by facilitating informed discussions with health care professionals.

## Discussion

This review and responsiveness evaluation of CMGs in MERS, SARS and COVID-19 demonstrates that, as was the case earlier in the pandemic,^1^ many CMGs have substantial methodological flaws and there has been little or no improvement between outbreaks/within the COVID-19 pandemic. The substantial heterogeneity observed in therapeutic recommendations at the beginning of the pandemic did however narrow as reliable evidence from clinical trials became available. The rationale for recommendations around supportive care was often unclear and the quality of evidence used to inform these was notably poor. Many CMGs recommended treatments despite them being non-evidence based or even having demonstrated futility. Despite a body of literature now available highlighting atypical presentations of COVID-19, particularly in elderly patients (e.g. less fever, more delirium, falls and diarrhoea^41^), and risk of more severe diseases, most guidelines did not provide specific advice for management of this patient group.^19 21 22 42-45^

### Unanswered questions and future research

Our review highlighted that recommendations on supportive care made by CMGs are often underpinned by limited and/or low-quality evidence. Where CMGs did conduct a systematic evidence review, this was usually primarily focussed on antiviral or immunomodulatory therapy. General aspects of supportive care (e.g. timing of intubation vs. a trial of NIV, target oxygen saturations, whether to give antibiotics, fluid balance decisions and thromboprophylaxis dose/agent/post-discharge regimen) are applicable to all viral infectious diseases with pandemic potential and especially important for emerging infections when the evidence base for pathogen specific therapy is limited. These issues should be addressed in living syndromic systematic reviews which would highlight knowledge gaps to be addressed in clinical trials and aid the rapid production of rigorous pathogen specific guidelines. Significant investment in the evidence base surrounding basic supportive care would likely yield great rewards in future and be globally applicable, especially given the relatively greater accessibility and lower cost of these interventions. At the onset of outbreaks, guideline committees could then identify pathogen specific clinical questions for which pragmatic RCTs could be established.

These results demonstrate the need for a better framework for the development of CMGs in outbreak settings. CMGs can still be useful and developed in a rigorous manner even when the quality and quantity of evidence available is minimal. Dissemination of expert opinion may be useful where there is no better option but should be clearly signposted as such and the rationale for recommendations needs to be clearly and transparently presented. We suggest that at least the initial methodology used to produce CMGs is subjected to a more transparent review process and ideally that these reviews should also be published. This is particularly pertinent given that the quality of CMGs did not appear to improve over time, and updated versions use a near identical format to the original. This need not slow their release which could initially be noted as interim guidance having not yet undergone such review (in a similar manner to preprints).

CMGs would benefit from incorporating succinct summaries, with decision making tools such as flowcharts and algorithms to aid rapid decision making on the front line. Patient groups should be involved in the development of CMGs from the beginning and lay summaries should be produced to enable patients to take a proactive and informed role in their care. Whilst this is more challenging in the initial phase of the pandemic, it would be feasible and desirable to have a pool of lay volunteers on standby who could be recruited at short notice to provide input into both guideline development and clinical research. As the COVID-19 pandemic has evolved, a variety of different issues have emerged, including atypical COVID-19 presentations,^41^ post COVID-19 syndrome^46^ and difficulty accessing medical care during lockdowns. Continuous engagement with all stakeholders would help to identify these issues and ensure that guidelines are responsive to them.

We observed substantial variation in the way that CMGs approach uncertainty when making recommendations on the basis of little and/or low-quality evidence. There are several examples where guidelines either recommended an unproven agent for use in particular patient groups or on a case-by-case basis. There is always a temptation for “compassionate use” of biologically plausible agents for individual patients *in extremis* with no proven treatment option.^47^ If however all patients who were treated with steroids/hydroxychloroquine/remdesivir/convalescent plasma had been randomised into trials from the beginning of the pandemic, we would have known whether these agents are beneficial (or indeed harmful) much sooner and more patients could have benefitted from these results. The success of pragmatic trials such as SOLIDARITY/RECOVERY have demonstrated the feasibility of this even in pandemic settings.^36 38^

### Strengths and weaknesses of the study

The inclusion of CMGs from a wide range of countries and organisations over a period of time is a strength of this study. This allowed us to evaluate the response of guideline committees to new emerging evidence. Our review is skewed towards countries in higher income classifications and we only identified one CMG from a low-income country (LIC).^48-50^ The AGREE-II tool is not specifically designed to appraise infectious disease CMGs produced during a pandemic, which may have caused us to underestimate the quality of some guidelines.

## Conclusions and policy implications

In conclusion, the quality of guidelines has not improved over time and despite publication of data from key clinical trials, some CMGs continue to recommend the use of agents found to be ineffective in RCTs. Existing guideline development frameworks which have successfully improved the quality of CMGs in general, have had minimal effect on those produced in response to epidemics and pandemics. This highlights a need for a CMG development framework for the production of timely, evidence based, resource conscious, locally adaptable and inclusive CMGs in response to emerging outbreaks. Vulnerable groups and in particular the elderly continue to be disproportionately overlooked and the relevant specialities (e.g. geriatrics) are underrepresented in CMG development groups. Given that COVID-19 has had such a profound impact on so many people’s lives and that such a vast quantity of public money has been spent, involvement of patients and the public in outbreak preparedness and response, including in CMG development is an area that needs to be urgently improved and must not be neglected in the future.

## Data Availability

Raw data has been deposited on Figshare.

https://doi.org/10.6084/m9.figshare.13561991.v1

## Authors’ Contributions

SL, IR and VC wrote the first draft of the manuscript, with input from LS and AD. EH performed the search strategy and executed the database search. SL performed the analysis of the AGREE-II scores and SL and IR created the figures. PB, EC, MT, TE, KL, LM, IR, SL, AD, MM, VC, and AVG screened the references, assisted with data and interpretation. KC provided additional comments from a lay perspective and helped to draft sections relating to PPI. LS and AD conceptualised the protocol and study. LS and PH provided overall supervision of the project. All authors reviewed and approved the final content for publication.

The corresponding author attests that all listed authors meet the authorship criteria and that no others meeting the criteria have been omitted.

## Ethical approval

We sought the opinion of the University of Oxford ethics committee who opined our involvement of a patient group constituted patient-public involvement and thus did not require ethical review.

## Data Sharing

Raw AGREE-II scores have been deposited on FigShare (https://doi.org/10.6084/m9.figshare.13561991.v1)

## Transparency statement

The lead author affirms that the manuscript is an honest, accurate and transparent account of the study being reported; no important aspects of the study have been omitted; any discrepancies from the study as originally planned have been explained.

## Acknowledgements

We would like to express our gratitude to the ISARIC network of Infectious disease physicians/public health practitioners who were valuable in searching and providing clinical management guidelines used in this review. Notably, the Ministry of Health Seychelles, Christine Williams, Dr. Desmond Oppong at the Greater Accra Regional Hospital, Ghana and Prof. Simon Anderson at the University of the West Indies. We thank our patient group for their suggestions and feedback. We thank the ISARIC global coordinating center for their help and support. Thanks to Sarah Dawson for providing advice regarding the formatting of the references. The authors also thank Julian Higgins for his critical reading of the manuscript.

## List of abbreviations

CMG: Clinical Management guidelines
SARS-CoV-1: Severe Acute Respiratory Syndrome Coronavirus-1
MERS-CoV: Middle East Respiratory Syndrome
Coronavirus COVID-19: Coronavirus Disease-19
SARS-CoV-2: Severe Acute Respiratory Syndrome Coronavirus-2
PREPARE: Platform for European Preparedness Against (Re-)emerging Epidemics
PROSPERO: International Prospective Register of Systematic Reviews
AGREE-II: Appraisal of Guidelines for Research and Evaluation II
HCID: High Consequence Infectious Disease
ISARIC: International Severe Acute Respiratory and emerging Infection Consortium
IQR: Interquartile Range
NIV: Non-invasive ventilation
IV: Intravenous
RECOVERY: Randomised Evaluation of COVID-19 Therapy
CDC: Centers for Disease Control and Prevention
IDSA: Infectious Diseases Society of America
WHO: World Health Organisation
REMAP-CAP: Randomised, Embedded, Multi-factorial, Adaptive Platform Trial for Community-Acquired Pneumonia
RCT: Randomised Control Trial
ACTT-1: Adaptive COVID-19 Treatment Trial
ARDS: Acute Respiratory Distress Syndrome

## References

1. Dagens A, Sigfrid L, Cai E, et al. Scope, quality, and inclusivity of clinical guidelines produced early in the covid-19 pandemic: rapid review. BMJ 2020;369:m1936. doi: 10.1136/bmj.m1936 [published Online First: 2020/05/28]

2. Williamson E, Walker AJ, Bhaskaran KJ, et al. OpenSAFELY: factors associated with COVID-19-related hospital death in the linked electronic health records of 17 million adult NHS patients. MedRxiv 2020 doi: 10.1101/2020.05.06.20092999

3. Public Health England. Disparities in the risk and outcomes of COVID-19, 2020.

4. Global Preparedness Monitoring Board. A World at Risk-Annual report on global preparedness for health emergencies: World Health Organization.

5. Moher D LA, Tetzlaff J, Altman DG, Group P. Preferred reporting items for systematic reviews and meta-analyses: the PRISMA statement. PLoS Med 2009 doi: doi: 10.1371/journal.pmed.1000097

6. International Severe Acute Respiratory and Emerging Infection Consortium. International Severe Acute Respiratory and emerging Infection Consortium 2020 [Available from: https://isaric.org/.

7. Graham R MM, Wolman D, Greenfield S, Steinberg E. Clinical guidelines we can trust. National Academies Press 2011;266 doi: DOI: 10.17226/13058

8. Brouwers MC KM, Browman GP, Burgers JS, Cluzeau F, Feder G, et al. AGREE II: advancing guideline development, reporting and evaluation in health care. Preventive Medicine 2010;51(5)

9. Hoffmann-Esser W, Siering U, Neugebauer EA, et al. Guideline appraisal with AGREE II: Systematic review of the current evidence on how users handle the 2 overall assessments. PLoS One 2017;12(3):e0174831. doi: 10.1371/journal.pone.0174831 [published Online First: 2017/03/31]

10. R Core Team. R: A language and environment for statistical computing,2017.

11. Wickham H. ggplot2: Elegant Graphics for Data Analysis: Springer-Verlag New York 2016.

12. The World Bank. The World Bank 2021 [Available from: https://www.worldbank.org/.

13. World Health Organisation. Clinical Management of COVID-19 2020 [updated 27 May 2020]. Available from: https://www.who.int/publications/i/item/clinical-management-of-covid-19 [Accessed: 10 June 2020].

14. Alhazzani Waleed MH, Møller; Yaseen, M. Arabi; Mark, Loeb; Michelle Ng, Gong; Eddy, Fan; Simon, Oczkowski; Mitchell, M. Levy; Lennie, Derde; Amy, Dzierba; Bin, Du; Michael, Aboodi; Hannah, Wunsch; Maurizio, Cecconi; Younsuck, Koh; Daniel, S. Chertow; Kathryn, Maitland; Fayez, Alshamsi; Emilie, Belley-Cote; Massimiliano, Greco; Matthew, Laundy; Jill, S. Morgan; Jozef, Kesecioglu; Allison, McGeer; Leonard, Mermel; Manoj, J. Mammen; Paul, E. Alexander; Amy, Arrington; John, E. Centofanti; Giuseppe, Citerio; Bandar, Baw; Ziad, A. Memish; Naomi, Hammond; Frederick, G. Hayden; Laura, Evans; Andrew, Rhodes. Surviving Sepsis Campaign: guidelines on the management of critically ill adults with Coronavirus Disease 2019 (COVID-19). Intensive Care Medicine 2020;46:854–87. doi: 10.1007/s00134-020-06022-5

15. World Health Organisation. Clinical management of severe acute respiratory infection when Middle East respiratory syndrome coronavirus (MERS-CoV) infection is suspected 2019 [updated January 2019]. Available from: https://www.who.int/csr/disease/coronavirus_infections/ipc-mers-cov/en/ [Accessed: 5 August 2020].

16. Yong Pil Cjy, Song; Yu Bin, Seo; JaePhil, Choi; HyoungShik, Shin; HeeJung, Yoon; JunYong, Choi; TaeHyong, Kim; YoungHwa, Choi; HongBin, Kim; JiHyun, Yoon; Jacob, Lee; JoongSik, Eom; JoonYoung, Song; SangOh, Lee; WonSup, Oh; HeeJin, Cheong; YoungGoo, Song; JungHyun, Choi; Woo Joo, Kim. Antiviral treatment guidelines for middle east respiratory syndrome. Infection and Chemotherapy 2015;47:212–22. doi: 10.3947/ic.2015.47.3.212

17. Adarsh BR, L. Morgan; Amy Hirsch, Shumaker; Valery, Lavergne; Lindsey, Baden; Vincent Chi-Chung, Cheng; Kathryn, M. Edwards; Rajesh, Gandhi; William, J. Muller; John, C. O., Shmuel, Shoham; M. Hassan Murad; Reem, A. Mustafa; Shahnaz, Sultan; Yngve, Falck-Ytter. Infectious Diseases Society of America Guidelines on the Treatment and Management of Patients with COVID-19 Version 1.0.3 2020 [updated 13 April 2020]. Available from: www.idsociety.org/COVID19guidelines. [ Accessed 4 November 2020].

18. Ministry of Health Family Welfare Bangladesh. National Guidelines on Clinical Management of Coronavirus Disease 2019 (Covid-19) Verison 4 2020 [updated 30 March 2020]. Available from: https://corona.gov.bd/documents/COVID_Guideline_V4.0..30.3.2020.pdf [Accessed: 2 June 2020].

19. Government of Pakistan Ministry of National Health Services. Clinical Management Guidelines for COVID-19 Infections Version 02 2020 [updated 1 June 2020]. Available from: https://www.nih.org.pk/wp-content/uploads/2020/06/20200106-Clinical-Management-Guidelines-for-COVID-19-infection-v2.pdf [Accessed 27 July 2020].

20. Министерство здравоохранения Российской. ВРЕМЕННЫЕ МЕТОДИЧЕСКИЕРЕКОМЕНДАЦИИПРОФИЛАКТИКА, ДИАГНОСТИКА И ЛЕЧЕНИЕ НОВОЙ КОРОНАВИРУСНОЙ ИНФЕКЦИИ (Covid-19) ВерсирF 7 (03.06.2020) 2020 [updated 3 June 2020]. Available from: https://static-0.rosminzdrav.ru/system/attachments/attaches/000/050/584/original/03062020_%D0%9CR_COVID-19_v7.pdf [Accessed: 27 July 2020].

21. Nepal Medical Council. Interim Clinical Guidance for Care of Patients with Covid-19 in Healthcare Settings 2020 [updated 3 April 2020]. Available from: https://nmc.org.np/files/4/NMC%20COVID-19%20Interim%20Clinical%20Guideline%20for%20care%203%20April.pdf [Accessed: 15 June 2020].

22. Tave F S; Michel R; Saha K B; Noel M; Hassanein M; Fayon M; Pugazhendhi S; Michel J; Pool B; Comarmond d L; Chetty A. Seychelles Clinical Guidelines for the Management of Severe Acute Respiratory Infection (SARI) in Patients with Confirmed COVID-19 Disease Version 1 2020 [updated 20 March 2020].

23. Ministry of Health Welfare Taiwan Centers for Disease Control. Interim Guidelines for Clinical Management of SARS-CoV-2 Infection 5^th^ Edition 2020 [updated 26 March 2020]. Available from: https://www.cdc.gov.tw/Uploads/d812c836-4956-4d4b-8775-a50e58e300b8.pdf [Accessed: 10 June 2020].

24. Haut Counseil de la Sante Publique. Avis relatif aux recommandations thérapeutiques dans la prise en charge du COVID-19(complémentaire à l’avis du 5 mars 2020) 2020 [updated 23 March 2020]. Available from: https://www.hcsp.fr/Explore.cgi/Telecharger?NomFichier=hcspa20200323_coronsarscovrecomthrap.pdf [Accessed: 22 June 2020].

25. Korean Society of Infectious Diseases. 신종코로나바이러스 검사에 대한 대한감염학회 권고안 2020 [updated 11 February 2020]. Available from: http://www.ksid.or.kr/eng/main/main.html [22 July 2020].

26. Centers for Disease Control Prevention. Interim Clinical Guidance for Management of Patients with Confirmed Coronavirus Disease (COVID-19) as of May 29, 2020 2020 [updated 29 May 2020]. Available from: https://www.cdc.gov/coronavirus/2019-ncov/hcp/clinical-guidance-management-patients.html [Accessed: 1 April 2020].

27. Adarsh BR, L. Morgan; Amy Hirsch, Shumaker; Valery, Lavergne; Lindsey, Baden; Vincent Chi-Chung, Cheng; Kathryn, M. Edwards; Rajesh, Gandhi; Jason, Gallagher; William, J. Muller; John, C. O., Shmuel, Shoham; M. Hassan Murad; Reem, A. Mustafa; Shahnaz, Sultan; Yngve, Falck-Ytter. Infectious Diseases Society of America Guidelines on the Treatment and Management of Patients with COVID-19 Verion 3.3 2020 [updated 25 June 2020]. Available from: www.idsociety.org/COVID19guidelines [Accessed: 4 November 2020].

28. Robert Koch Institute. Hinweise zu Erkennung, Diagnostik und Therapie von Patienten mit COVID-19 Stand: März 2020 2020 [updated March 2020]. Available from: https://www.bmj.com/lookup/google-scholar?link_type=googlescholar&gs_type=article&q_txt=.+Diagnosis+and+treatment+of+patiens+with+COVID-19.Robert+Koch+Institute%2C+2020. [Accessed: 6 June 2020].

29. Robert Koch Institute. Hinweise zu Erkennung, Diagnostik und Therapie von Patienten mit COVID-19 Stand: 09.10.2020 2020 [updated 9 October 2020]. Available from: www.stakob.rki.de https://www.rki.de/DE/Content/Kommissionen/Stakob/Stellungnahmen/Stellungnahme-Covid-19_Therapie_Diagnose.pdf?blob=publicationFile [ Accessed: 15 October 2020].

30. Министерство здравоохранения Российской. ВРЕМЕННЫЕ МЕТОДИЧЕСКИЕ РЕКОМЕНДАЦИИ ПРОФИЛАКТИКА, ДИАГНОСТИКА И ЛЕЧЕНИЕ НОВОЙ КОРОНАВИРУСНОЙ ИНФЕКЦИИ (COVID-19) Версия 8 (03.09.2020) 2020 [updated 3 September 2020]. Available from: https://static-0.minzdrav.gov.ru/system/attachments/attaches/000/051/777/original/030902020_COVID-19_v8.pdf?utm_source=yxnews&utm_medium=desktop [Accessed: 1 October 2020].

31. Centers for Disease Control Prevention. Interim Clinical Guidance for Management of Patients with Confirmed Coronavirus Disease (COVID-19) as of September 10, 2020 2020 [updated 10 September 2020]. Available from: https://www.cdc.gov/coronavirus/2019-ncov/hcp/clinical-guidance-management-patients.html [10 September 2020].

32. 衛生福利部疾病管制署. 新型冠 病毒(SARS-CoV-2)感染臨床處置暫行指引 2020 [updated 17 August 2020]. Available from: https://www.cdc.gov.tw/ [Accessed: 1 September 2020].

33. Darmon M ME, Morawiec E, Schnell D, Maury E, Constantin M J, Montravers P et al. Recommandations d’experts portant sur la prise en charge en réanimation des patients infectés à SARS-CoV2 Version 5 du 07/11/2020 2020 [updated 7 November 2020]. Available from: https://sfar.org/download/recommandations-dexperts-portant-sur-la-prise-en-charge-en-reanimation-des-patients-en-periode-depidemie-a-sars-cov2/ [Accessed: 17 November 2020].

34. Coreb Mission Nationale. Recommandations d’experts portant sur la prise en charge en réanimation des patients en période d’épidémie à SARS-CoV2 Version 4 2020 [updated 7 April 2020. Available from: https://sfar.org/recommandations-dexperts-portant-sur-la-prise-en-charge-en-reanimation-des-patients-en-periode-depidemie-a-sars-cov2/ [Accessed: 10 July 2020].

35. Darmon MB, Lila; Morawiec, Elise; Maury, Eric; Constantin, Jean-Michel; Montravers, Philippe; Zahar, Jean-Ralph; Lucet, Jean-Christophe; Guery, Benoit; Bessis, Simon; Saidani, Nadia; Desmettre, Thibault; Dumas, Florence;. Recommandations d’experts portant sur la prise en charge en réanimation des patients en période d’épidémie à SARS-CoV2 Version 2 du 10/03/2020 2020 [updated 10 March 2020]. Available from: https://www.srlf.org/wp-content/uploads/2020/03/Recommandations-dexperts-COVID-19-10-Mars-2020.pdf [Acessed: 16 March 2020].

36. The RECOVERY Collaborative Group. Dexamethasone in Hospitalized Patients with Covid-19 — Preliminary Report. The New England Journal of Medicine 2020 doi: DOI: 10.1056/NEJMoa2021436

37. The Writing Committee for the REMAP-CAP Investigators. Effect of Hydrocortisone on Mortality and Organ Support in Patients With Severe COVID-19: The REMAP-CAP COVID-19 Corticosteroid Domain Randomized Clinical Trial. JAMA 2020;324(13):1317–29. doi: 10.1001/jama.2020.17022

38. WHO Solidarity Trial Consortium. Repurposed Antiviral Drugs for Covid-19 — Interim WHO Solidarity Trial Results. New England Journal of Medicine 2020 doi: 10.1056/NEJMoa2023184

39. Beigel H J Tmk, Dodd E Lori et al., for the ACTT-1 study Group Members. Remdesivir for the Treatment of Covid-19 — Final Report. New England Journal of Medicine 2020;383:1813–26. doi: DOI: 10.1056/NEJMoa2007764

40. Stockman LJ, Bellamy R, Garner P. SARS: systematic review of treatment effects. PLoS Med 2006;3(9):e343. doi: 10.1371/journal.pmed.0030343 [published Online First: 2006/09/14]

41. Kerr AD, Stacpoole SR. Coronavirus in the elderly: a late lockdown UK cohort. Clin Med (Lond) 2020;20(6):e222–e28. doi: 10.7861/clinmed.2020-0423 [published Online First: 2020/09/12]

42. Ministry of Health Cuba. Infecciones por coronavirus – COVID-19 2020 [Available from: https://temas.sld.cu/coronavirus/covid-19/ [Acessed: 5 June 2020].

43. Ghana Ministry of Health. Provisional Standard Treatment Guidelines for Novel Coronavirus Infection COVID -19 Guidelines for Ghana Version 1.0 2020 [Available from: https://www.moh.gov.gh/wp-content/uploads/2016/02/COVID-19-STG-JUNE-2020-1.pdf [Accessed: 20 June 2020].

44. Pakistan Chest Society. COVID-19 Management Guidelines 2020 2020 [updated 28 March 2020]. Available from: http://www.pakistanchestsociety.pk/wp-content/uploads/2020/03/COVID-19-Management-guideline-PCS-28-March.pdf [Accessed: 27 August 2020].

45. Department of Health Republic of South Africa. Clinical management of suspected or confirmed COVID-19 disease Version 3 2020 [updated 27 March 2020]. Available from: https://www.nicd.ac.za/wp-content/uploads/2020/03/Clinical-Management-of-COVID-19-disease_Version-3_27March2020.pdf [Accessed: 28 June 2020].

46. National Health Services. Post-COVID Syndrome (Long COVID) 2020 [Available from: https://www.england.nhs.uk/coronavirus/post-covid-syndrome-long-covid/.

47. Rojek AM, Martin GE, Horby PW. Compassionate drug (mis)use during pandemics: lessons for COVID-19 from 2009. BMC Med 2020;18(1):265. doi: 10.1186/s12916-020-01732-5 [published Online First: 2020/08/23]

48. Ministerio Da Saude. Suspeito de COVID-19 2020 [updated March 2020].

49. Ministério da Saúde. OrientaÇÕes Para Manejo De Pacientes Com Covid-19 2020 [Available from: https://portalarquivos.saude.gov.br/images/pdf/2020/June/18/Covid19-Orientac--o--esManejoPacientes.pdf [Acessed: 1 July 2020].

50. Ministério Da Saúde. Fluxograma de Pacientes com Suspeita de Covid-19 nos Serviços ambulatórios (Triagem e Consultas Externas) 2020 [Available from: http://www.misau.gov.mz/index.php/normas-procedimentos-e-fluxos?download=239:fluxogramas-e-protocolos [Accessed: 24 July 2020].

51. European Respiratory Society. COVID-19: Guidelines and recommendations directory 2020 [Available from: https://www.ersnet.org/covid-19-guidelines-and-recommendations-directory.

52. Ministerio de Salud Argentina. Recomendaciones Para El Abordaje Terapeutico Version 2 2020 [updated 29 May 2020]. Available from: https://www.argentina.gob.ar/salud [Accessed: 1 June 2020].

53. National Covid-Clinical Evidence Taskforce. Australian Guideline for the Clinical Care of People with COVID-19 2020 [Available from: https://covid19evidence.net.au/ [Accessed:20 July 2020].

54. Ministry of Public Health. Directives Et Procedures Operationnelles Standards Pour La Preparation Et La Reponse Au Covid-19 Au Cameroun 2020 [updated April 2020].

55. Ministry of Health Cuba. Infecciones por coronavirus – SARS [Available from: https://temas.sld.cu/coronavirus/sars-cov/ [Accessed: 5 June 2020].

56. Ministry of Health Cuba. Infecciones por coronavirus – MERS [Available from: https://temas.sld.cu/coronavirus/mers/ [Accessed: 5 June 2020].

57. National Institute for Research in Reproductive Health Indian Council of Medical Research. Guidance for Management of Pregnant Women in COVID-19 Pandemic 2020 [Available from: http://www.nirrh.res.in/ [Accessed: 14 June 2020].

58. Stefan K, Janssens U, Welte T, Weber-Carstens S, Marx G, Karagiannidis C,. S1-Empfehlungen zur intensivmedizinischen Therapie von Patienten mit COVID-19 Version 1 2020 [updated March 2020]; cited 123 123]. Available from: https://link.springer.com/article/10.1007/s00063-020-00674-3 [Accessed on 2 November 2020]46.

59. Ministry of Health Family Welfare India. Revised Guidelines on Clinical Management of COVID – 19 2020 [updated 31 March 2020]. Available from: https://www.mohfw.gov.in/pdf/RevisedNationalClinicalManagementGuidelineforCOVID1931032020.pdf [Accessed: 2 June 2020].

60. Ministero della Salute. Gestione clinica dell ‘ infezione respiratoria acuta grave nei casi di sospetta infezione da nuovo coronavirus (nCoV) 2020 [updated 12 January 2020]. Available from: http://www.salute.gov.it/imgs/C_17_pagineAree_5373_5_file.pdf [Accessed: 24 July 2020].

61. Jamaica Ministry of Health and Wellness. Guideline for the management of Pregnancy during the COVID-19 Pandemic 2020 [updated 25 March 2020]. Available from: https://www.moh.gov.jm/covid-19-resources-and-protocols/ [17 August 2020].

62. Jamaica Ministry of Health and Wellness. Covid-19 Preparedness and Response Plan for Outbreak Control Clinical Management of Severe Acute Respiratory Infection When Novel Coronavirus Covid-19 Infection Is Suspected Version 2 2020 [updated March 2020]. Available from: www.moh.gov.jm https://www.moh.gov.jm/wp-content/uploads/2020/04/2019nCoV-COVID-19-Clinical-Management-Guidelines-V2.pdf [Accessed: 17 August 2020].

63. 厚生労働省. 新型コロナウイルス感染症 Covid-19 V2 2020 [updated 18 May 2020]. Available from: https://www.mhlw.go.jp/index.html [Accessed: 27 July 2020].

64. 厚生労働省. 新型コロナウイルス感染症(COVID-19) 診療の手引き_V3 2020 [updated 3 September 2020]. Available from: https://www.mhlw.go.jp/content/000670444.pdfhttps://www.mhlw.go.jp/stf/seisakunitsuite/bunya/0000121431_00111.html [Accessed: 9 September 2020].

65. Chicamba Valeria LS, ; Nhamtubo Carmita, et al. Orientações para a Abordagem e Tratamento do doente Pediátrico na UCIP do Hospital Central Maputo com COVID-19 2020 [

66. Nigeria Centre for Disease Control. National Interim Guidelines for Clinical Management of COVID-19 Version 1 2020 [updated 14 March 2020]. Available from: https://ncdc.gov.ng/themes/common/docs/protocols/177_1584210847.pdf [Accessed: 27 June 2020].

67. National Committee for Management of Covid-United Arab Emirates. National Guidelines for Clinical Management and Treatment of COVID-19 Version 2 2020 [updated 3 April 2020]. Available from: https://www.dha.gov.ae/en/HealthRegulation/Documents/COVID%20National%20Guidelines%20FINAL%2018%20March.pdf [Accessed: 20 May 2020].

68. National Health Service. Clinical management of persons admitted to hospital with suspected COVID-19 infection Version 1 2020 [updated 19 March 2020]. Available from: https://www.england.nhs.uk/coronavirus/wp-content/uploads/sites/52/2020/03/clinical-management-of-persons-admitted-to-hospita-v1-19-march-2020.pdf [Accessed: 24 August 2020].

69. Maria Laura Vc, Siroti; Guillermo, Montiel; Ada, Toledo; Carlos, Franceschini; Alejandro. Martinez, Fraga; Leslie. Vargas, Ramirez; JoseLuis, Carillo; Martha. Torres, Fraga. Recomendaciones para el Manejo No Invasivo e Invasivo de la Insuficiencia Respiratoria Hipoxémica de Novo Covid--19. 2020 [Available from: https://www.aamr.org.ar/secciones/coronavirus/recomendaciones_soporte_ventilatorio_ovid.pdfc [23 July 2020].

70. World Health Organisation. Clinical management of severe acute respiratory infection 2020 [updated March 2020]. Available from: https://www.who.int/docs/default-source/coronaviruse/clinical-management-of-novel-cov.pdf [Accessed: 1 April 2020].

71. The Korean Society of Pediatric Infection Diseases. COVID-19 Guidelines: Pediatric Care 2020 [updated 20 March 2020]. 44]. Available from: http://www.ksid.or.kr/eng/main/main.html [Accessed: 21 July 2020].

72. Korean Society for Infectious Diseases Korean Society for Antibacterial Therapy Korean Society for Pediatric Infections Korean Society for Tuberculosis; Respiratory System. 코로나 19 (COVID-19) 약물 치료에 관한 전문가 권고안 (version 1.1) 2020 [updated 25 February 2020]. Available from: http://www.ksid.or.kr/eng/main/main.html [Accessed: 22 July 2020].

73. 이무식. 시론 지역사회와 함께 하는 코로나 19 (Covid-19) 극복 V 1.1 2020 [updated 1 March 2020]. Available from: http://www.ksid.or.kr/eng/main/main.html [Accessed: 1 June 2020].

74. Lim S W Ars, Read C R, on behalf of the SARS guidelines committee of the British Thoracic Society,. Hospital management of adults with severe acute respiratory syndrome (SARS) if SARS re-emerges -updated 10 February 2004. Journal of Infection 2004;49:1–7. doi: 10.1016/j.jinf.2004.04.001

75. National Health Comission of the People’s Republic of China. 中东呼吸综合征病例诊疗方案(2015年版) 2015 [updated 6 June 2015]. Available from: http://en.nhc.gov.cn/ [Accessed: 27 July 2020].

76. National Health Comission of the People’s Republic of China. 传染性非典型肺炎(Sars)诊疗方案(2004版) 2005 [updated 25 May 2005]. Available from: http://en.nhc.gov.cn/ [Accessed: 27 July 2020].

77. National Health Comission of the People’s Republic of China. 新型冠状病毒肺炎诊疗方案 (试行第八版) 2020 [updated 19 August 2020]. Available from: http://en.nhc.gov.cn/ [Accessed: 17 September 2020].

78. Chinese Medical Association CAoCM. Consensus of the management of severe acute respiratory syndrome. Zhonghua yi xue za zhi 2003;83:1731–52.

79. De La Famille Et Des Personnes Handicapees Ministere De La Sante. Conduite à tenir pour la prise en charge des personnes présentant un syndrome ou une 2004 [updated 06 April 2004]. Available from: www.sante.gouv.fr http://umvf.omsk-osma.ru/infectiologie/www.infectiologie.com/site/medias/alertes/sars/protoc_060404.pdf [Accessed: 22 June 2020].

80. Maxwell C MA, Tai KF, Seimer A,. SOGC Clinical Practice Guideline. Management guidelines for obstetric patients and neonates born to mothers with suspected or probable severe acute respiratory syndrome (SARS). No. 225, April 2009. International journal of gynaecology and obstetrics: the official organ of the International Federation of Gynaecology and Obstetrics 2009;107:82–86. doi: 10.1016/j.ijgo.2009.05.006

81. National Health Commission. Diagnosis and Treatment Protocol for Novel Coronavirus Pneumonia (Trial Version 7). Chinese Medical Journal 2020;133 doi: 10.1097/CM9.0000000000000819

82. Ministerio De Sanidad. Documento técnico Manejo clínico de pacientes con enfermedad por el nuevo coronavirus (COVID-19) 2020 [updated 3 March 2020]. Available from: https://www.seipweb.es/wp-content/uploads/2020/03/Protocolo_manejo_clinico_COVID-19.pdf [Accessed: 10 March 2020].

83. Ministerio de Sanidad. Manejo clínico del COVID-19: unidades de cuidados intensivos 2020 [updated 18 June 2020]. Available from: https://www.mscbs.gob.es/en/home.htm [Accessed: 25 June 2020].

84. Rijksinstituut voor Volksgezondheid en Milieu. Medicamenteuze behandeling voor patiënten met COVID-19 (infectie met SARS–CoV-2) 2020 [updated 13 August 2020] Available from: https://swab.nl/nl/covid-19 [Accessed: 9 September 2020].

85. Società Italiana di Malattie Infettive e Tropicali. Linee Guida Sulla Gestione Terapeutica e di Supporto per Pazienti con Infezione da Coronavirus COVID-19 2020 [updated March 2020]. Available from: http://www.fvcalabria.unicz.it/COVID-19/LINEE-GUIDA/linee-guida-SIMIT-marzo-2020.pdf [1 April 2020].

86. Vollaard AE, Pauline; Gieling, Emilie; Boer de, Mark; Snijder, Erik; Dissel Van, Jaap. Medicamenteuze behandelopties voor opgenomen patiënten met COVID-19 2020 [updated 3 March 2020]. Available from: https://www.rivm.nl/ [Accessed: 1 April 2020].

87. Ministerio De Sanidad. Documento técnico Manejo clínico del COVID-19: atención hospitalaria 2020 [updated 18 June 2020]. Available from: https://www.mscbs.gob.es/profesionales/saludPublica/ccayes/alertasActual/nCov/documentos/Protocolo_manejo_clinico_ah_COVID-19.pdf [25 June 2020].

88. British Thoracic Society and Scottish Intercollegiate Guidelines Network. SIGN158 British guideline on the management of asthma. A national clinical guideline. 2003 [updated July 2019. Available from: https://www.brit-thoracic.org.uk/quality-improvement/guidelines/asthma2020.

89. Società Italiana Malattie Infettive e Tropicali Sezione Regione Lazio. Gruppo di Lavoro COVID-19-SIMIT Lazio Comitato di Redazione 2020 [updated 5 May 2020]. Available from: https://www.simi.it/ [Acessed: 10 May 2020].

90. Ministerio De Sandidad. Documento técnico Manejo de la mujer embarazada y el recién nacido con COVID-19 2020 [updated 13 May 2020]. Available from: https://www.mscbs.gob.es/en/home.htm [Accessed: 1 June 2020].

