## Supplement for "From SARS and MERS to COVID-19: a review of the quality and responsiveness of clinical management guidelines in outbreak settings"

Search Strategy

**Grey literature search strategy for individual countries Ministry of Health and National Public Health websites, and for searching the other organisations (e.g. US CDC etc.)**

**Search Terms:**

**MERS-CoV**

MERS-CoV clinical management guidelines

MERS-CoV guidelines

Strategies for management of MERS-CoV

Clinical practice guidelines for management of MERS-CoV

Clinical management of MERS-CoV

**SARS**

SARS clinical management guidelines,

SARS guidelines

Strategies for management of SARS

Clinical practice guidelines for management of SARS

Clinical management of SARS

Severe Acute Respiratory Infections clinical management guidelines,

Severe Acute Respiratory Infections clinical guidelines

Strategies for clinical management of Severe Acute Respiratory Infections

Clinical practice guidelines for management of Severe Acute Respiratory Infections

Clinical management of Severe Acute Respiratory Infections

**COVID-19**

COVID-19 clinical management guidelines,

COVID-19 clinical guidelines

Strategies for clinical management of COVID-19

Clinical practice guidelines for management of COVID-19

Clinical management of COVID-19

Novel Coronavirus clinical management guidelines,

Novel Coronavirus clinical guidelines

Strategies for clinical management of novel coronavirus

Clinical practice guidelines for management of novel coronavirus

Clinical management of novel coronavirus

**Google search**

Translated using google translate to language (or languages) used in country

Searched google region specific search (Settings -> Search Settings -> Region Settings) on google.

First 50 entries examined

Additionally, specifically search Ministry of Health/Public Health agency websites if not found in google search.

**If not found using above methods document that no guideline was identified via the grey literature search. Subsequent attempts to acquire the guideline via local contacts will be made.**

Data extraction form

| **Assessment group** | **Questions** | **Answer format type** |
| --- | --- | --- |
| Availability | Name of the guideline | Short-answer text |
|  | First three author names (in the form Full surname initial of first name) | Short-answer text |
|  | Organisation | Short-answer text |
|  | Year of latest revision | Day, Month, Year |
|  | Type of geography | Multiple Choice (Worldwide, Regional (specific to a continent, country specific) |
|  | Geographical aim (e.g., China, UK, USA) | Short-answer text |
|  | Disease(s)/syndrome(s) covered | Multiple Choice (COVID-19, SARS-CoV, MERS-CoV) |
| Inclusivity | Populations covered | Multiple Choice (Children, Pregnant Women, HIV/Immunocompromised, Older people, Adults, Not specified) |
|  | If children - are/is there a specific age range(s) given? | Long-answer text |
|  | Other details on population (e.g., specific age groups etc.) | Long-answer text |
| Scope | Is a case definition provided? | Multiple Choice (Yes, No) |
| Therapeutic/Supportive care recommendations | Are antibiotics recommended? | Multiple Choice (Yes, No) |
|  | Describe the antibiotic recommendation by population | Long-answer text |
|  | What is the supporting evidence for antibiotic recommendations? (Links to scientific literature, expert consensus, links to another guideline etc.) | Long-answer text |
|  | Are antivirals recommended? | Multiple Choice (Yes, No) |
|  | Which specific antivirals are recommended? | Long-answer text |
|  | Describe antiviral recommendation by population | Long-answer text |
|  | What is the supporting evidence for antiviral recommendations? (Links to scientific literature, expert consensus, links to another guideline etc.) | Long-answer text |
|  | Are corticosteroids recommended? | Multiple Choice (Yes, No) |
|  | Describe corticosteroid recommendation by population | Long-answer text |
|  | What is the supporting evidence for corticosteroid recommendations? (Links to scientific literature, expert consensus, links to another guideline etc.) | Long-answer text |
|  | Is fluid therapy recommended? | Multiple Choice (Yes, No) |
|  | What is the supporting evidence for fluid therapy recommendations? (Links to scientific literature, expert consensus, links to another guideline etc.) | Long-answer text |
|  | Is oxygen therapy recommended? | Multiple Choice (Yes, No) |
|  | Do oxygen therapy recommendations differ by population? (if yes - how?) | Long-answer text |
|  | What is the supporting evidence for oxygen therapy? (Links to scientific literature, expert consensus, links to another guideline etc.) | Long-answer text |
|  | Are any other treatments recommended? (if yes - which?) | Long-answer text |
|  | Are any other treatments recommended? (if yes - which?) | Long-answer text |
|  | What is the supporting evidence for any other recommendations? (Links to scientific literature, expert consensus, links to another guideline etc.) | Long-answer text |
|  | Other comments | Long-answer text |

Guidelines included in this review

| **Guideline** | **Guidelines included in this review** | | | | |
| --- | --- | --- | --- | --- | --- |
|  | **Reference** | **Syndrome** | **Date Published** | **Version** | **Authorising body** |
| **Argentina** | **52** | COVID-19 | May-20 | 2 | Ministerio de Salud |
| **Argentina** | **69** | COVID-19 |  |  | Argentina Association of Respiratory Medicine |
| **Australia** | **53** | COVID-19 |  |  | Australian National COVID-19 Clinical Evidence Taskforce |
| **Bangladesh** | **18** | COVID-19 | Mar-20 | 4 | Ministry of Health and Family Welfare |
| **Brazil** | **49** | COVID-19 |  |  | Ministerio da Saúde |
| **British Thoracic Society** | **74** | SARS | Feb-04 |  | W. S. Lim, S. R. Anderson, R. C. Read et al |
| **Cameroon** | **54** | COVID-19 | Apr-20 | 1 | Ministère de la Santé Publique |
| **Canadian Obstetrics Society** | **80** | SARS | Apr-09 |  | C. Maxwell, A. McGeer, K. F. Tai, A. Seimer |
| **China** | **81** | COVID-19 | Mar-20 | 7 | National Health Commission & National Administration of Traditional Chinese Medicine |
| **China** | **77** | COVID-19 | Aug-20 | 8 | National Health Comission of the People's Republic of China |
| **China** | **75** | MERS | Jun-15 |  | National Health Comission of the People's Republic of China |
| **China** | **76** | SARS | May-05 |  | National Health Comission of the People's Republic of China |
| **Chinese Medical Association** | **78** | SARS | Jun-05 |  | N. Zhong, Y. Ding, Y. Mao, Q. Wang, G. Wang, D. Wang, Y. Cong, Q. Li, Y. Liu, L. Ruan, B. Chen, X. Du, Y. Yang, Z. Zhang, X. Zhang, J. Lin, J. Zheng, Q. Zhu, D. Ni, X. Xi, G. Zeng, D. Ma, C. Wang, W. Wang, B. Wang, J. Wang |
| **Cuba** | **55** | SARS |  |  | Ministerio De Salud Pública |
| **Cuba** | **56** | MERS |  |  | Ministerio De Salud Pública |
| **Cuba** | **42** | COVID-19 |  |  | Ministerio De Salud Pública |
| **France** | **79** | SARS | 6-Apr-04 |  | Ministere De La Sante De La Famille Et Des Personnes Handicapees |
| **France** | **24** | COVID-19 | Mar-20 |  | Haut Counseil de la Sante Publique |
| **France (COREB)** | **34** | COVID-19 | Apr-20 | 2 | Darmon M, Bouadma L, Morawiec E et al., |
| **France (COREB)** | **35** | COVID-19 | Mar-20 | 4 | Coreb Mission Nationale |
| **France (COREB)** | **33** | COVID-19 | Nov-20 | 5 | Darmon M, Bouadma L, Morawiec E et al., |
| **Germany** | **58** | COVID-19 | Mar-20 | 1 | Kluge S, Janssens U, Welte T et al. |
| **Ghana** | **43** | COVID-19 |  | 1 | Ghana Ministry of Health |
| **India** | **59** | COVID-19 | Mar-20 |  | Ministry of Health & Family Welfare |
| **India** | **57** | COVID-19 |  |  | ICMR - National Institute for Research in Reproductive Health |
| **Infectious Diseases Society of America** | **27** | COVID-19 | Jun-20 |  | Adarsh Bhimraj, Morgan R, Hirsch Shumaker A, et al. |
| **Infectious Diseases Society of America** | **17** | COVID-19 | Apr-20 |  | Adarsh Bhimraj, Morgan R, Hirsch Shumaker A, et al. |
| **Italy** | **60** | COVID-19 | Jan-20 |  | Ministero della Salute |
| **Italy, Società Italiana di Malattie Infettive e Tropicali,** | **85** | COVID-19 | Mar-20 |  | Società Italiana di Malattie Infettive e Tropicali |
| **Italy, Società Italiana di Malattie Infettive e Tropicali,** | **89** | COVID-19 | May-20 |  | COVID-19 –SIMIT Lazio Working Group |
| **Jamaica** | **61** | COVID-19 | Mar-20 | 2 | Jamaica Ministry of Health and Wellness |
| **Jamaica** | **62** | COVID-19 | Mar-20 |  | Jamaica Ministry of Health and Wellness |
| **Japan** | **63** | COVID-19 | May-20 | 2 | Ministry of Health, Labor and Welfare |
| **Japan** | **64** | COVID-19 | Sep-20 | 3 | Ministry of Health, Labor and Welfare |
| **Korea Society of Infectious Diseases** | **16** | MERS |  |  | Yong Pil, Chong; Joon Young, Song; Yu Bin, Seo; Jae Phil, Choi; Hyoung Shik, Shin; Hee Jung, Yoon; Jun Yong, Choi; Tae Hyong, Kim; Young Hwa, Choi; Hong Bin, Kim; Ji Hyun, Yoon; Jacob, Lee; Joong Sik, Eom; Joon Young, Song; Sang Oh, Lee; Won Sup, Oh; Hee Jin, Cheong; Young Goo, Song; Jung Hyun, Choi; Woo Joo, Kim |
| **Mozambique** | **65** | COVID-19 | Mar-20 |  | Ministério Da Saúde |
| **Mozambique** | **48** | COVID-19 | Mar-20 |  | Ministério Da Saúde |
| **Mozambique** | **50** | COVID-19 | Mar-20 |  | Ministério Da Saúde |
| **Nepal** | **21** | COVID-19 | Apr-20 |  | Nepal Medical Council COVID-19 Treatment Guidance Committee |
| **Nigeria (Nigeria CDC)** | **66** | COVID-19 | Mar-20 | 1 | Nigeria Centre for Disease Control |
| **Pakistan** | **19** | COVID-19 | Jun-20 | 2 | Government of Pakistan Ministry of National Health Services |
| **Pakistan** | **44** | COVID-19 | Mar-20 |  | Pakistan Chest Society |
| **Rijksinstituut voor Volksgezondheid en Milieu** | **86** | COVID-19 | Mar-20 |  | Vollaard A, Ellerbroek P, Gieling E et al., |
| **Rijksinstituut voor Volksgezondheid en Milieu** | **84** | COVID-19 | Aug-20 |  | Vollaard A, Ellerbroek P, Gieling E et al., |
| **Robert Koch Institute** | **29** | COVID-19 | Oct-20 |  | Feldt T, Guggemos W, Heim K et al., Robert Koch Institute |
| **Robert Koch Institute** | **28** | COVID-19 | Mar-2020 |  | Robert Koch Institute |
| **Russia** | **20** | COVID-19 | Jun-20 | 7 | Министерство здравоохранения Российской Федерации |
| **Russia** | **30** | COVID-19 | Sep-20 | 8 | ВРЕМЕННЫЕ МЕТОДИЧЕСКИЕ РЕКОМЕНДАЦИИ ПРОФИЛАКТИКА, ДИАГНОСТИКА И ЛЕЧЕНИЕ НОВОЙ КОРОНАВИРУСНОЙ ИНФЕКЦИИ (COVID-19) Версия 8 (03.09.2020) |
| **Seychelles** | **22** | COVID-19 | Mar-20 | 1 | Tave Susan F, Michel R, Saha Barun K et al., |
| **South Africa** | **45** | COVID-19 | Mar-20 | 3 | Health Department Republic of South Africa, National Institute for Communicable Diseases |
| **South Korea** | **71** | COVID-19 | Mar-20 |  | The Korean Society of Pediatric Infection Diseases |
| **South Korea** | **72** | COVID-19 | Feb-20 |  | Korean Society for Infectious Diseases Korean Society for Antibacterial Therapy Korean Society for Pediatric Infections Korean Society for Tuberculosis; Respiratory System, |
| **South Korea** | **73** | COVID-19 | Mar-20 | 1.1 | The Korean Society for Critical Care, The Korean Society of Tuberculosis and Respiratory System The Korean Society for Antibacterial Therapy Severe Corona 19 Infection (COVID-19) Treatment System |
| **South Korea** | **25** | COVID-19 | Feb-20 |  | Korean Society of Infectious Diseases |
| **Spain** | **87** | COVID-19 | Jun-2020 |  | Ministerio De Sanidad |
| **Spain** | **83** | COVID-19 | Jun-2020 |  | Ministerio De Sanidad |
| **Spain** | **82** | COVID-19 | Mar-2020 |  | Ministerio De Sanidad |
| **Spain** | **90** | COVID-19 | May-2020 |  | Ministerio De Sanidad |
| **Surviving Sepsis Campaign** | **14** | COVID-19 | Mar-20 |  | Waleed A, Morten H M, Yaseen M A et al., |
| **Taiwan (Taiwan CDC)** | **23** | COVID-19 | Mar-20 | 5 | Ministry of Health and WelfareTaiwan Centres for Disease Control |
| **Taiwan (Taiwan CDC)** | **32** | COVID-19 | Aug-20 |  | Ministry of Health and WelfareTaiwan Centres for Disease Control |
| **United Arab Emirates** | **67** | COVID-19 | Apr-20 | 2 | National committee for Management of COVID-19 Cases |
| **United Kingdom** | **68** | COVID-19 | Mar-20 | 1 | National Health Service England |
| **United States (CDC)** | **31** | COVID-19 | Sep-20 |  | Centers for Disease Control and Prevention |
| **United States (CDC)** | **26** | COVID-19 | Apr-20 |  | Centers for Disease Control and Prevention |
| **WHO** | **70** | COVID-19 | Mar-20 | 2 | World Health Organisation |
| **WHO** | **13** | COVID-19 | May-20 |  | World Health Organisation |
| **WHO** | **15** | MERS | Oct-19 |  | World Health Organisation |

Key:

Merged guidelines- grey

Different versions of the same guideline- Blue

AGREE-II domain and average scores

| **Guideline** | **AGREE-II domain scores of each guideline included in this review** | | | | | | | | |
| --- | --- | --- | --- | --- | --- | --- | --- | --- | --- |
|  | **Reference** | **Scope and purpose (%)** | **Stake Holder Involvement (%)** | **Rigor of Development (%)** | **Clarity of Presentation (%)** | **Applicability (%)** | **Editorial Indpend-ence (%)** | **Average domain scores (%)** | **Average overall score** |
| **Argentina** | 52 | 19.44 | 11.11 | 28.13 | 55.56 | 39.58 | 8.33 | 27.03 | 4 |
| **Argentina (Association of Respiratory Medicine)** | 69 | 30.56 | 16.67 | 17.71 | 44.44 | 37.50 | 0.00 | 24.48 | 3.5 |
| **Australia** | 53 | 69.44 | 52.78 | 77.08 | 88.89 | 62.50 | 41.67 | 65.39 | 5.5 |
| **Bangladesh** | 18 | 33.33 | 33.33 | 28.13 | 52.78 | 43.75 | 0.00 | 31.89 | 3.5 |
| **Brazil** | 49 | 69.44 | 41.67 | 13.54 | 44.44 | 35.42 | 0.00 | 34.09 | 3.5 |
| **British Thoracic Society** | 74 | 50.00 | 27.78 | 23.96 | 72.22 | 27.08 | 0.00 | 33.51 | 4.5 |
| **Cameroon** | 54 | 75.00 | 72.22 | 0.00 | 63.89 | 41.67 | 12.50 | 44.21 | 3 |
| **Canadian Obstetrics Society** | 80 | 63.89 | 30.56 | 37.50 | 52.78 | 22.92 | 4.17 | 35.30 | 3.5 |
| **China** | 81 | 33.33 | 8.33 | 6.25 | 69.44 | 14.58 | 0.00 | 21.99 | 3.5 |
| **China** | 77 |  |  |  |  |  |  |  |  |
| **China** | 75 | 16.67 | 5.56 | 6.25 | 30.56 | 8.33 | 0.00 | 11.23 | 2 |
| **China** | 76 | 16.67 | 5.56 | 4.17 | 25.00 | 20.83 | 0.00 | 12.04 | 3 |
| **Chinese Medical Association** | 78 | 58.33 | 41.67 | 35.42 | 75.00 | 29.17 | 0.00 | 39.93 | 4.5 |
| **Cuba** | 55 | 19.44 | 2.78 | 7.29 | 19.44 | 12.50 | 0.00 | 10.24 | 2 |
| **Cuba** | 56 | 16.67 | 8.33 | 11.46 | 2.78 | 0.00 | 0.00 | 6.54 | 1.5 |
| **Cuba** | 42 | 41.67 | 19.44 | 7.29 | 2.78 | 16.67 | 0.00 | 14.64 | 2 |
| **France** | 79 | 30.56 | 11.11 | 10.42 | 25.00 | 27.08 | 8.33 | 18.75 | 2 |
| **France** | 24 | 30.56 | 33.33 | 27.08 | 47.22 | 27.08 | 4.17 | 28.24 | 4 |
| **France (COREB)** | 34 | 55.56 | 36.11 | 31.25 | 55.56 | 27.08 | 66.67 | 45.37 | 4.5 |
| **France (COREB)** | 35 |  |  |  |  |  |  |  |  |
| **France (COREB)** | 33 |  |  |  |  |  |  |  |  |
| **Germany** | 58 | 30.56 | 27.78 | 20.83 | 30.56 | 27.08 | 4.17 | 23.50 | 2 |
| **Ghana** | 43 | 77.78 | 36.11 | 23.96 | 83.33 | 35.42 | 0.00 | 42.77 | 4.5 |
| **India** | 59 | 83.33 | 33.33 | 50.00 | 80.56 | 62.50 | 0.00 | 51.62 | 5.5 |
| **India** | 57 |  |  |  |  |  |  |  |  |
| **Infectious Diseases Society of America** | 27 |  |  |  |  |  |  |  |  |
| **Infectious Diseases Society of America** | 17 | 75.00 | 66.67 | 88.54 | 100.00 | 45.83 | 83.33 | 76.56 | 6 |
| **Italy** | 60 | 63.89 | 36.11 | 38.54 | 66.67 | 47.92 | 4.17 | 42.88 | 4.5 |
| **Italy, Società Italiana di Malattie Infettive e Tropicali,** | 85 | 41.67 | 13.89 | 55.21 | 66.67 | 14.58 | 30 | 37.00 | 4 |
| **Italy, Società Italiana di Malattie Infettive e Tropicali,** | 89 | 41.67 | 27.78 | 57.29 | 91.67 | 29.17 | 12.50 | 43.34 | 4 |
| **Jamaica** | 61 | 80.56 | 38.89 | 28.13 | 94.44 | 60.42 | 0.00 | 50.41 | 5.5 |
| **Jamaica** | 62 |  |  |  |  |  |  |  |  |
| **Japan** | 63 | 30.56 | 36.11 | 12.50 | 47.22 | 16.67 | 20.83 | 27.31 | 3.5 |
| **Japan** | 64 |  |  |  |  |  |  |  |  |
| **Korea Society of Infectious Diseases** | 16 | 19.44 | 16.67 | 19.79 | 66.67 | 31.25 | 0.00 | 25.64 | 3 |
| **Mozambique** | 65 | 41.67 | 11.11 | 12.50 | 47.22 | 31.25 | 8.33 | 25.35 | 2.5 |
| **Mozambique** | 48 |  |  |  |  |  |  |  |  |
| **Mozambique** | 50 |  |  |  |  |  |  |  |  |
| **Nepal** | 21 | 80.56 | 36.11 | 17.71 | 72.22 | 37.50 | 0.00 | 40.68 | 5 |
| **Nigeria** | 66 | 50.00 | 22.22 | 9.38 | 66.67 | 22.92 | 0.00 | 28.53 | 3.5 |
| **Pakistan** | 19 | 33.33 | 27.78 | 16.67 | 69.44 | 20.83 | 0.00 | 28.01 | 4.5 |
| **Pakistan Chest Society** | 44 | 41.67 | 55.56 | 16.67 | 58.33 | 27.08 | 0.00 | 33.22 | 3.5 |
| **Rijksinstituut voor Volksgezondheid en Milieu** | 86 | 33.33 | 16.67 | 76.04 | 77.78 | 12.50 | 12.50 | 38.14 | 4 |
| **Rijksinstituut voor Volksgezondheid en Milieu** | 84 | 61.11 | 52.78 | 45.83 | 63.89 | 39.58 | 8.33 | 45.25 | 4.5 |
| **Robert Koch Institute** | 29 | 61.11 | 30.56 | 27.08 | 50.00 | 37.50 | 8.33 | 35.76 | 3.5 |
| **Robert Koch Institute** | 28 | 58.33 | 27.78 | 10.42 | 61.11 | 4.17 | 12.50 | 29.05 | 4 |
| **Russia** | 20 | 75.00 | 47.22 | 9.38 | 69.44 | 22.92 | 12.50 | 39.41 | 3 |
| **Russia** | 30 | 69.40 | 58.30 | 31.00 | 69.40 | 77.10 | 12.50 | 52.95 | 4.5 |
| **Seychelles** | 22 | 86.11 | 58.33 | 28.13 | 88.89 | 25.00 | 8.33 | 49.13 | 5.5 |
| **South Africa** | 45 | 47.22 | 19.44 | 25.00 | 66.67 | 29.17 | 0.00 | 31.25 | 4 |
| **South Korea** | 71 | 94.44 | 55.56 | 97.92 | 97.22 | 70.83 | 100.00 | 86.00 | 6.5 |
| **South Korea** | 72 | 44.44 | 25.00 | 18.75 | 69.44 | 14.58 | 0.00 | 28.70 |  |
| **South Korea** | 73 |  |  |  |  |  |  |  |  |
| **South Korea** | 25 |  |  |  |  |  |  |  |  |
| **Spain** | 82 | 72.22 | 47.22 | 23.96 | 75 | 37.50 | 5 | 43.48 | 6 |
| **Spain** | 90 |  |  |  |  |  |  |  |  |
| **Spain** | 83 | 80.60 | 37.50 | 26.20 | 88.90 | 35.40 | 0.00 | 44.80 | 5 |
| **Spain** | 87 | 75.00 | 36.11 | 17.71 | 83.33 | 18.75 | 16.67 | 41.26 | 4 |
| **Surviving Sepsis Campaign** | 14 | 80.56 | 63.89 | 86.46 | 97.22 | 68.75 | 100.00 | 82.81 | 6 |
| **Taiwan (Taiwan CDC)** | 23 | 44.44 | 11.11 | 28.13 | 66.67 | 25.00 | 0.00 | 29.22 | 5 |
| **Taiwan (Taiwan CDC)** | 32 |  |  |  |  |  |  |  |  |
| **United Arab Emirates** | 67 | 58.33 | 30.56 | 22.92 | 52.78 | 33.33 | 4.17 | 33.68 | 4 |
| **United Kingdom (National Health Service)** | 68 | 44.44 | 19.44 | 19.79 | 50.00 | 20.83 | 0.00 | 25.75 | 3 |
| **United States (CDC)** | 31 | 88.89 | 36.11 | 47.92 | 86.11 | 50.00 | 45.83 | 59.14 | 5 |
| **United States (CDC)** | 26 | 63.89 | 27.78 | 38.54 | 63.89 | 18.75 | 12.50 | 37.56 | 4 |
| **WHO** | 70 | 55.56 | 27.78 | 64.58 | 75.00 | 12.50 | 17.50 | 42.40 | 4 |
| **WHO** | 13 | 86.11 | 55.56 | 45.83 | 86.11 | 33.33 | 66.67 | 62.27 | 5.5 |
| **WHO** | 15 | 77.78 | 50.00 | 36.46 | 97.22 | 41.67 | 33.33 | 56.08 | 5.5 |

Key:

Merged guidelines- grey

Different versions of the same guideline- Blue
